# Discrepancies between registered protocol and final publication in exercise interventions for chronic low back pain: a meta-research study

**DOI:** 10.1101/2023.02.27.23286399

**Authors:** Silvia Bargeri, Giacomo Basso, Ignazio Geraci, Greta Castellini, Alessandro Chiarotto, Silvia Gianola, Raymond Ostelo, Marco Testa, Tiziano Innocenti

**Author notes:** co-first authors. Alphabetic order except for co-first, second and last authors.

## Abstract

**Background:** Evidence of selective reporting bias is common in randomized controlled trials (RCTs) of various medical fields, undermining their integrity and credibility. However, this has not yet been assessed in exercise for chronic low back pain (cLBP) RCTs. Therefore, we will aim to assess the prevalence of discrepancies between the registered protocol and final publication in this field and the characteristics of RCTs with and without such discrepancies.

**Methods:** We will start from the RCTs included in the 2021 Cochrane review (n=249) and identified in its update (n=172) to select all RCTs reporting a protocol registration. Standardized data collection form will be developed to record information from both registration and publication. We will then detect discrepancies for primary and secondary outcomes, outcomes measures, timepoints, number of arms and statistical analysis plans between the registered protocol and final publication. We will use descriptive statistics to assess the proportion of RCTs with and without a discrepancy as well as to compare their characteristics.

**Ethics and dissemination:** We will offer insights and recommendations for future RCTs avoiding selective reporting that can reflect in subsequent inaccuracies in systematic reviews or guidelines for clinical practice. Results of this study will be shared through conference presentations and publication in a peer-reviewed journals.

## 1. Introduction

Low back pain is one the greatest contributors to years lived with disability and is the first cause of activity limitation, and absence from work ^7 21^. One widely used intervention for chronic low back pain (CLBP) is exercise therapy, which has been examined in numerous randomized controlled trials (RCTs)^6^. Due to their important effect on clinical practice, there is a necessity to have transparent reporting of RCT results^9^. However, bias in the design, conduct or reporting of RCTs can result in inaccuracies in systematic reviews or guidelines and subsequent errors in clinical practice^4^.

Several meta-research studies in the medical field^2-4, 10, 19, 22^ have shown that discrepancies between what is reported in the registered protocol and what is reported in the final publication are common. This can lead to selective reporting bias and refers to a publication practice where study authors preferentially publish interesting or positive research findings while concealing results that do not confirm their hypothesis because of the statistical significance, magnitude or direction of the effect^14 11 15^. Despite some improvement over time, it has been shown that the study quality and reporting of trials in the exercise for CLBP field continue to be lacking^5^. However, it is still unclear what is the prevalence of discrepancies between the registered protocol and final publication in these trials. This could strongly affect the conclusions of systematic review, overestimating the effects of an intervention or underestimating its undesirable effect, compromising the credibility of the evidence synthesis itself.

Starting from the largest updated Cochrane review on the effectiveness of exercise intervention in CLBP^6^ we will aim to assess:

- The prevalence of RCTs with a discrepancy between the registered protocol and final publication in primary and secondary outcomes according to outcomes measures, timepoints, number of arms, and statistical analysis plans
- The characteristics of RCTs with and without discrepancies between the registered protocol and final publication

## 2. Methods

### Study design

We will conduct a meta-research study ^17, 18^. Since the reporting checklist for methods research studies is currently under development ^12^, we will adapt items from the Preferred Reporting Items for Systematic Reviews and Meta-analyses (PRISMA) for reporting meta-research studies ^13^.

### Eligibility criteria

We will start from the RCTs included in the 2021 Cochrane review”^6^ (n=249) and identified in its update (n=172) to select all RCTs reporting a protocol registration. Hayden et al. included RCTs that compared exercise to no treatment, usual care, placebo or another conservative intervention among adults with CLBP. Trials could include interventions provided to participants in any setting (e.g., healthcare, occupational, general and mixed populations). The intervention could have been combined with or without the addition of other components (eg, education, manual therapy).

Protocols will be considered when they were registered to a primary register of the WHO International Clinical Trials Registry Platform (ICTRP) or in ClinicalTrials.gov according to the ICMJE8. If no information about protocol registration is reported, if the protocol is not available or not in English language, we will exclude it.

### Data extraction

Standardised data collection forms will be used to record information from registered protocol and the final publication of the trial.

For data extraction of the registered protocol, we will collect: registration date, study start date, primary outcome registration date, primary completion date (i.e., date of final collection of data for the primary outcome), registered number of arms, description of interventions, statistical analysis plans, nature and number of primary and secondary outcomes (e.g, pain), time points (e.g., 1 month follow up) and outcome measurements (e.g., visual analogue scale). We will also collect how many version of the registered protocol exists.

For data extraction of the final publication, we will use the dataset of the Cochrane review to extract RCTs’ general characteristics (e.g., author, year, ID number of the protocol registration and/or reference of protocol publication, initial date of participant enrollment, setting, sample size, CLBP duration (e.g., months), radicular symptoms (e.g. leg pain and/or neurological symptoms), mean age, sex, conflict of interests, funding (non-industry/industry-sponsored), journal of publication, journal impact factor (JIF), number of arms, description of interventions, statistical analysis, nature and number of primary and secondary outcomes, time points and outcome measurements).

We will classify the trial status into 1) prospectively registered; 2) retrospectively registered according to its registration date. Prospective registration will be defined as trial registration before or within a month of the first participant enrollment start date according to the protocol^1^.

### Detection of discrepancies between registered protocol and final publication

We will define discrepancies as differences between registered protocol (i.e., from the last prospectively registered version released) and final publication. To ensure a comprehensive assessment, we will check related documents for each RCT (e.g., published protocol, statistical analysis plans, supplementary materials).

Two pairs of two independent reviewers (SB, GB; IG, SG) will detect discrepancies for primary and secondary outcomes, outcomes measures, time points, number of arms and statistical analysis plans. We will adapt a previously published method16 to classify discrepancies into: change in definition (e.g., outcome proposed) or measure (e.g., VAS instead of NPRS), addition (e.g., completely outcome measure or arm added, new timepoint added), omission (e.g., excluded primary outcome, excluded arm). In case of switching between primary and secondary outcome we will classify it into upgrade (secondary outcome changed to primary) and downgrade (primary outcome changed to secondary). We will also collect if discrepancies between the original registered protocol and the last registered version are present with the corresponding date.

If no primary outcome will be explicitly defined within the manuscript, we will consider the outcome used for the power calculation to be the primary published outcome. In case of multiple outcomes/time points are planned in the registered protocol, but not reported in the final publication, we will check related publications referring to the same protocol.

We will distinguish between discrepancies reported and not reported in the final publication (i.e., deviation transparently reported in the manuscript), checking the final publication for an explanation of any deviation from the protocol. If deviations are transparently declared and likely to be justified, we will not consider them as discrepancies.

Before starting the assessment, a calibration phase will be performed by the four reviewers (SB, GB, IG, SG) piloting a small sample of 4 RCTs with protocols posted in different registries. Disagreements will be discussed during a debrief meeting with another reviewer (GC) to reach a final consensus.

### Comparison between discrepant outcomes and statistically significant results

According to a previous study^20^, a discrepancy will be considered to favour statistically significant results when: 1) a non-statistically significant (p-value > 0.05 or a confidence interval that crossed zero for continuous outcomes) primary outcome registered in the protocol will be downgraded to a secondary in the final publication; 2) a statistically significant secondary outcome registered in the protocol will be upgraded to a primary outcome in the final publication; and 3) addition of a non-registered statistically significant primary outcome in the final publication. We will prioritise results of between-group comparisons. If more time points are available, we will collect any comparison favouring the exercise intervention. If between-groups comparison is not available, we will collect within-group results favouring the exercise group versus control.

### Statistical analysis

We will use descriptive statistics to assess the proportion of RCTs with and without a discrepancy between the registered protocol and final publication as well as to compare their characteristics (e.g., funding received industry-sponsored/non industry sponsored, sample size, JIF, prospective/retrospective registration, published protocol yes/no). Data analysis will be performed with STATA software.

## Data Availability

All data produced in the present work are contained in the manuscript

## ETHICS AND DISSEMINATION

This study does not require an ethics review as we will not collect personal data; it will summarise information from publicly available studies. A manuscript will be prepared and submitted for publication in an appropriate peer-reviewed journal. The study findings will be disseminated at national and international conferences in research methods and musculoskeletal rehabilitation. We will offer insights and recommendations for future research and practice avoiding selective reporting that can reflect in subsequent inaccuracies in systematic reviews or guidelines for clinical practice.

## AUTHOR CONTRIBUTIONS

TI, SB, GB, IG conceived and designed the study protocol. SB, GB, IG, GC, AC, SG, RO, TI were involved in conceptualising the study objectives, providing input into study selection criteria and plans for data extraction. All the authors, including SB, GB, IG, GC, AC, SG, RO, MT, TI approved the final version of the protocol.

